# Markov computational modeling to predict opioid vs. money choice and behavioral effort in regular heroin users

**DOI:** 10.64898/2026.08.18.26360767

**Authors:** Amolak S. Jhand, Mark K. Greenwald

**Affiliations:** Dept. of Psychiatry and Behavioral Neurosciences, School of Medicine, Wayne State University, Detroit, MI, USA

**Keywords:** Opioid, Choice, Seeking, Effort, Computational modeling

## Abstract

Quantifying decision-making in experimental settings that mimic real-world conditions may provide insights into mechanisms underlying addiction. This study developed a computational model of opioid-seeking behavior. Out-of-treatment persons who regularly used heroin were stabilized on buprenorphine 8mg/day to minimize opioid withdrawal. Across programmatically-linked studies, three experimental conditions presented differing money vs. opioid unit amounts that could be earned per trial ($2 vs. 1-mg hydromorphone, *n*=23; $2 vs. 2-mg hydromorphone, *n*=36; $4 vs. 2-mg hydromorphone, *n*=24), controlling other factors. Progressive ratio schedules on each choice option required increasing effort across trials to earn the same amount. Trial-level outcomes were decision latency and choice on each option, and session-level outcomes were drug-money latency and breakpoint difference scores. A Markov computational model was used to predict the probability of choosing the same option as the previous trial (vs. switching). Model inputs included ‘effort discrepancy’ (between earning the same vs. other commodity on next choice) and logarithm of the ratio of decisional speed (current vs. previous choice). Participants who more rapidly chose hydromorphone vs. money made more consecutive drug choices and expended greater effort earning hydromorphone. First-trial hydromorphone choice predicted continued effortful opioid-seeking. Participants repeated choices on 80% of trials; the model accurately predicted ‘stick vs. switch’ behavior on 93% of trials. Participants typically repeated choices when faced with lower effort discrepancies and higher hydromorphone dose (2-mg vs. 1-mg). In conclusion, a Markov computational model accurately predicted effortful behavior in a choice paradigm that mimics real-world decisions between opioid and nondrug reinforcers.

## Introduction

Opioid misuse remains a public health crisis in North America. Despite recent decreases in opioid overdose deaths in the United States, approximately 74,000 Americans died by opioid overdose in the past year [1], and psychostimulants (primarily methamphetamine and cocaine) now contribute substantively to overdose deaths [2, 3]. Current FDA-approved medications for opioid use disorder (MOUD) include the full and partial μ-opioid agonists methadone and buprenorphine, which are often combined with psychosocial and behavioral therapies [4, 5].

Strong evidence indicates that MOUD reduces overdose and mortality [6–10], yet only ≈25% of US adults with OUD receive MOUD [11]. Even among patients who receive MOUD and are retained in treatment, many continue to use illicit opioids, especially those who inject, use synthetic opioids, or use psychostimulants [12–17]. Furthermore, the majority return to using opioids within the first year post-treatment [18–20]. To improve outcomes and reduce harms from OUD, it is critical to advance scientific understanding of mechanisms that contribute to drug-seeking and use behaviors.

Computational modeling is a valuable approach to formalize processes, and to estimate the dynamics of, drug-seeking behavior. In practice, ‘computational modeling’ in the field of addiction refers to a broad umbrella of methodologies. First, it is important to recognize that models/algorithms have been applied at different scales and for varying purposes (e.g. [21]) ranging from molar-level prediction of population trends (e.g. [22–24]); to meso-level analysis of an individual’s habitual vs. goal-directed behavioral states [e.g. 25–27], drug craving and its connection to use (e.g. [28–30]), or to allostasis or neuroadaptations that occur over longer time periods (e.g. [31, 32]); as well as micro-level context-dependent decisions and behaviors (e.g. [33, 34]), which is the topic of this paper. Second, many theoretical accounts of addiction have focused on reinforcement learning typically as it relates to acquisition of drug use or initial transition to addiction (e.g. [35–37]), which is less relevant for studying clinically important questions of persistent drug use and how this might be ameliorated. Although laboratory animal studies have increasingly studied factors that affect ecologically-meaningful drug vs. nondrug choices, few have investigated trial-by-trial sequences or within-session-level patterns using computational modeling of drug choice/seeking behavior (e.g. [38–40]). We presently lack human data on actual (i.e. reinforced, rather than hypothetical) choice and seeking of concurrently-available drug vs. non-drug reinforcers.

Several reviews have focused on proposed various mechanistic explanations of processes that contribute to drug use [41–46]. Among these, the behavioral economic approach offers a computational model of person- and session-level behavioral choice and price-sensitivity (micro-economic demand curve analysis) that relates to addiction severity [47]. A complementary approach, “pico-economic” analysis [48, 49], has been used to examine competing reinforcer value on a more ‘molecular’ time scale (trial-by-trial choice), but to our knowledge there are no relevant empirical data in human drug users. In real-world situations, people who use drugs repeatedly face choices to pursue vs. forego drug use, and choice parameters in a given context can strongly influence propensity for repetitive drug use [43, 45, 50]. Such ecologically-relevant scenarios can be simulated and mathematically modeled in the controlled laboratory setting.

Borrowing from micro- and pico-economic frameworks, the present study developed a computational model of opioid-seeking behavior. This analysis incorporated behavioral *choice* and *effortful response* data from participants who were out-of-treatment regular heroin users, gathered across four programmatically linked studies with prospectively-designed parametric variations while holding constant other factors. These studies varied (1) absolute and relative magnitudes of opioid (hydromorphone dose) vs. monetary choice options, and (2) required behavioral effort across within-session choice trials (progressive ratio schedule with independent exponential response ladders, which maximizes differences in relative effort as the participant persists on the same option). In all these studies, participants were stabilized on a fixed, moderate dose of buprenorphine that effectively suppressed opioid withdrawal symptoms (to minimize the likelihood that participants responded for drug simply to alleviate withdrawal).

Under these conditions, we examined how experimental parameters (hydromorphone and money unit amounts) and individual difference factors (e.g. sex, recent cocaine use, first-trial choice responding) influenced drug vs. money choice latencies and effortful drug responding.

We further developed a computational model to predict trial-by-trial choice persistence vs. switching as a function of differential effort and successive choice latency discrepancy. Fitted model parameters were compared across experimental conditions and participant subgroups, including first-trial choice and recent cocaine use. We hypothesized that, in this population of out-of-treatment chronic regular heroin users, we would observe a mixture of *habitual* and *goal-directed* behavior [27, 39, 40–45]; specifically, within-session trial-by-trial choice patterns would be partly habitual (i.e. same-choice sequences accompanied by shorter decision latencies reflecting less deliberation) but also partly goal-directed in being sensitive to variation of unit opioid dose and monetary reinforcement (i.e. disruption of same-choice chains in the presence of greater choice effort discrepancy and longer decision latencies reflecting greater deliberation).

Accordingly, we view drug choice/seeking as overlearned but malleable behavior, inconsistent with a purely compulsive behavioral perspective. Finally, we hypothesized that certain individual differences (e.g. first-trial drug choice, pre-experimental cocaine use) might influence choice patterns.

## Materials and methods

The Wayne State University IRB approved all procedures for the 4 source studies for this secondary data analysis. Each study was registered on ClinicalTrials.gov (NCT00218309 [IRB# 080403M1F], NCT00218361 [IRB# 063305MP2F], NCT00608504 [IRB# 062107MP2F], and NCT00684840 [IRB# 118407MP4F]). All studies were conducted in compliance with ethical standards outlined in the Belmont Report and Declaration of Helsinki. Each participant provided written and witnessed informed consent, separately for screening and their respective study.

Overall methods were similar across these studies, and have been previously described, so are summarized briefly here. De-identified data from these studies were initially compiled for analysis in March 2024 and the analyses were completed in May 2026.

### Participants

Male and female volunteers from 18–55 years of age, self-identifying as regular heroin users and not presently seeking treatment for their problematic opioid use, were recruited from the Detroit/metropolitan area using advertisements and word-of-mouth referral. Candidates were screened using medical history, routine blood and urine chemistry, electrocardiogram, tuberculin test, physical exam, and psychiatric interview (SCID-IV [51]). Volunteers were excluded who met DSM-IV criteria for current Axis I disorders (except Opioid and Nicotine Dependence), serious psychiatric conditions (psychosis, bipolar, major depression not substance-induced), reported chronic health problems or taking prescribed medications, were cognitively impaired (estimated verbal IQ < 80) based on the Shipley Institute of Living Scale [52]), or scored >15 on the 10-item Injection and Blood Withdrawal Phobia subscale of the Medical Fear Survey [53].

Women who were pregnant (urine HCG), lactating or heterosexually active and not using medically-approved birth control (self-report) were excluded; to improve feasibility of recruiting female participants, hormonal birth control was allowed.

During screening, participants completed the Drug History and Use Questionnaire (available on request), which provides a comprehensive lifetime and current assessment of substance use. Volunteers had to provide a urine sample positive for opioids (>300 ng/ml) and negative for methadone (due to planned buprenorphine maintenance in these studies), benzodiazepines (<300 ng/ml) and barbiturates (<200 ng/ml). Urine samples positive for cocaine (>300 ng/ml) or THC (>50 ng/ml) were allowed. Volunteers had to provide an alcohol-free breath sample (<.02%). The studies were conducted prior to widespread adulteration of the heroin supply with synthetic opioids and other substances (e.g. xylazine). Thus, exclusions for urine drug screen results were likely based on intentional use and not a contaminated supply.

Participants were classified based on screening data as either recent cocaine users (Coc+), defined as testing urinalysis-positive for cocaine or self-reporting any past-month cocaine use; or non-users (Coc–) if they tested cocaine urine-negative and self-reported no past-month cocaine use.

### Protocol timeline

Participants were stabilized on buprenorphine 8-mg/day sublingual for ≥10 outpatient days before inpatient admission. Residential living, staff observation and daily urinalyses ensured abstinence from unsanctioned drug use during study procedures. Testing was conducted on consecutive weekdays. Residential living, staff observation and daily urine-drug and breath-alcohol testing ensured abstinence from unsanctioned substance use during test sessions. During non-experimental periods (evenings and weekends), volunteers could engage in recreational activities on the unit. Cigarette smoking or use of nicotine replacement products (provided by research staff) were allowed *ad libitum* on the residential unit and, on session days, permitted until starting the drug/money choice task and resumed after the choice task.

### Drug administration

#### Buprenorphine

Participants consumed buprenorphine 8-mg sublingual tablets during initial outpatient and inpatient periods (Subutex™; supplied by Research Triangle Institute, Research Triangle Park, NC). During each experimental (inpatient) study, buprenorphine was always administered at 8:00 pm. The 8-mg/day buprenorphine dose was selected to minimize opioid withdrawal symptoms, but insufficient to block reinforcing effects of hydromorphone [54, 55]. This was intentionally designed so that participants were not engaging in opioid seeking simply to alleviate withdrawal, i.e. we sought to minimize the potential contribution of negative reinforcement motivation, such that positive reinforcement mechanisms were more likely to guide behavior. Following the completion of each study, participants were provided with a standardized three-week outpatient buprenorphine dose taper (not relevant to this analysis).

#### Hydromorphone

Doses of hydromorphone (HYD; Dilaudid-HP™ supplied in 50-mg/5-mL ampoules) were injected into the deltoid muscle. Maximum earned doses were either 12-mg (1.2 mL) or 24-mg (2.4 mL), as described below.

### Experimental conditions

By design, several shared experimental conditions were included across these four programmatically linked studies [56–59]. This enabled us to include participants across studies in the present analysis, improving our sample size and power for testing hypotheses. We identified three experimental conditions for our analysis (summarized in **Table 1**):

**Table 1.**
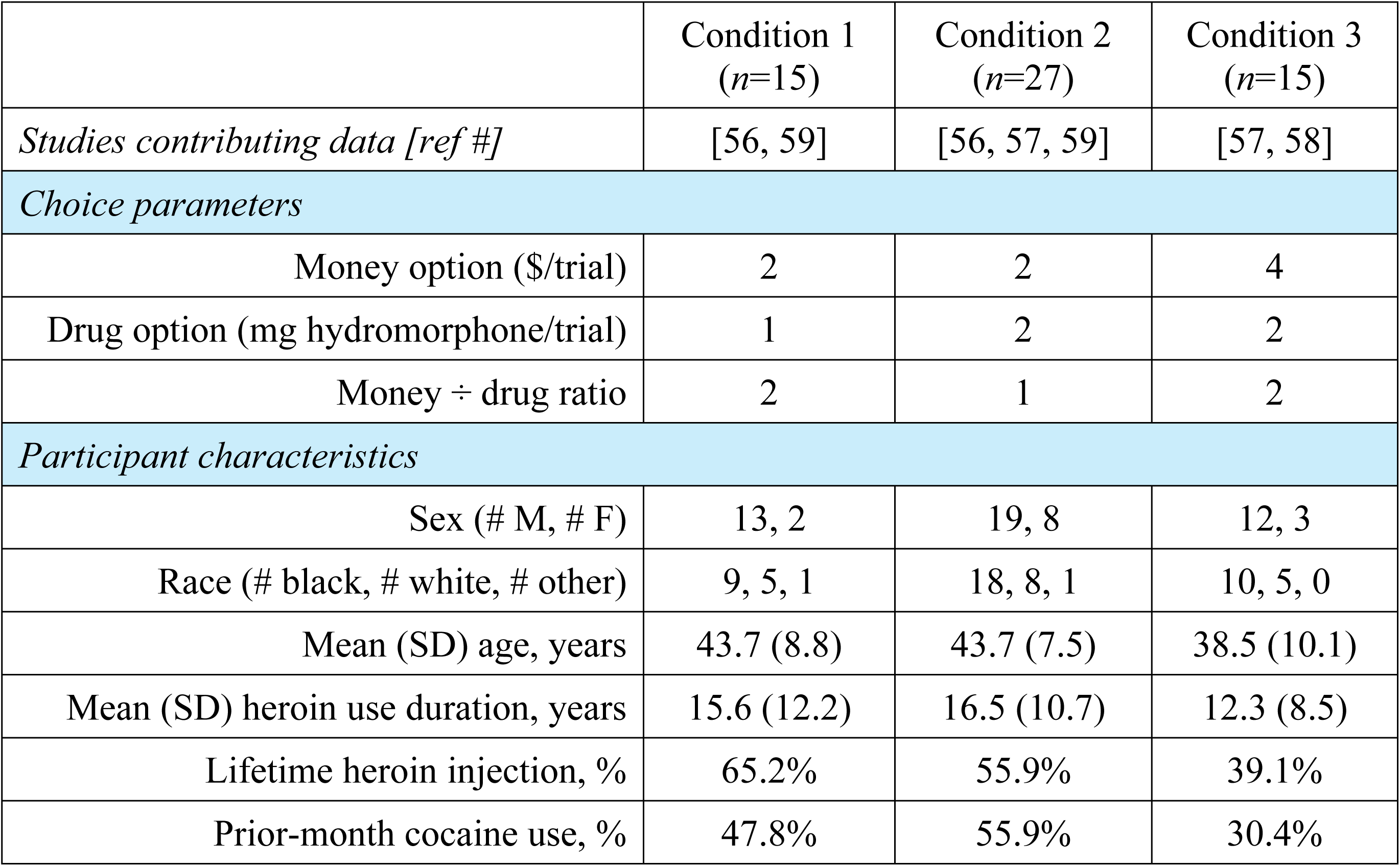
Choice parameters and participant characteristics in each experimental condition.

<u>Condition 1</u> includes 23 participants from [56] and [59] given the opportunity to choose on each trial between earning $2 and 1-mg HYD units in the absence of any session pretreatment (i.e. no supplemental HYD condition in [56] and placebo yohimbine condition in [59]).

<u>Condition 2</u> includes 36 participants from [56], [57], and [59] given the opportunity to choose on each trial between earning $2 and 2-mg HYD units in the absence of any session pretreatment or post-treatment (i.e. no supplemental HYD condition in [56] and [57], and placebo yohimbine condition in [59]).

Condition 3 includes 24 participants from [57] and [58] given the opportunity to choose on each trial between earning $4 and 2-mg HYD units in the absence of any session pretreatment.

### Choice session structure

During each experimental drug/money choice session, participants could work towards the drug and money reward amounts specified by the condition. A total of 12 choices between drug and money were possible during each 3-hr session. After choosing the commodity on each trial (one computer mouse click, FR1), the participant then had to work (click the mouse many more times) to earn that commodity. The behavioral effort or ‘price’ required to earn that same commodity again on subsequent choice trials during the session increased exponentially on a progressive ratio schedule, independently for drug and money (**Fig 1, left panel**). The amount of HYD earned was injected as a bolus intramuscularly by a nurse at the end of each session, and the amount of money earned was added to the participant’s study payment.

**Fig 1.**
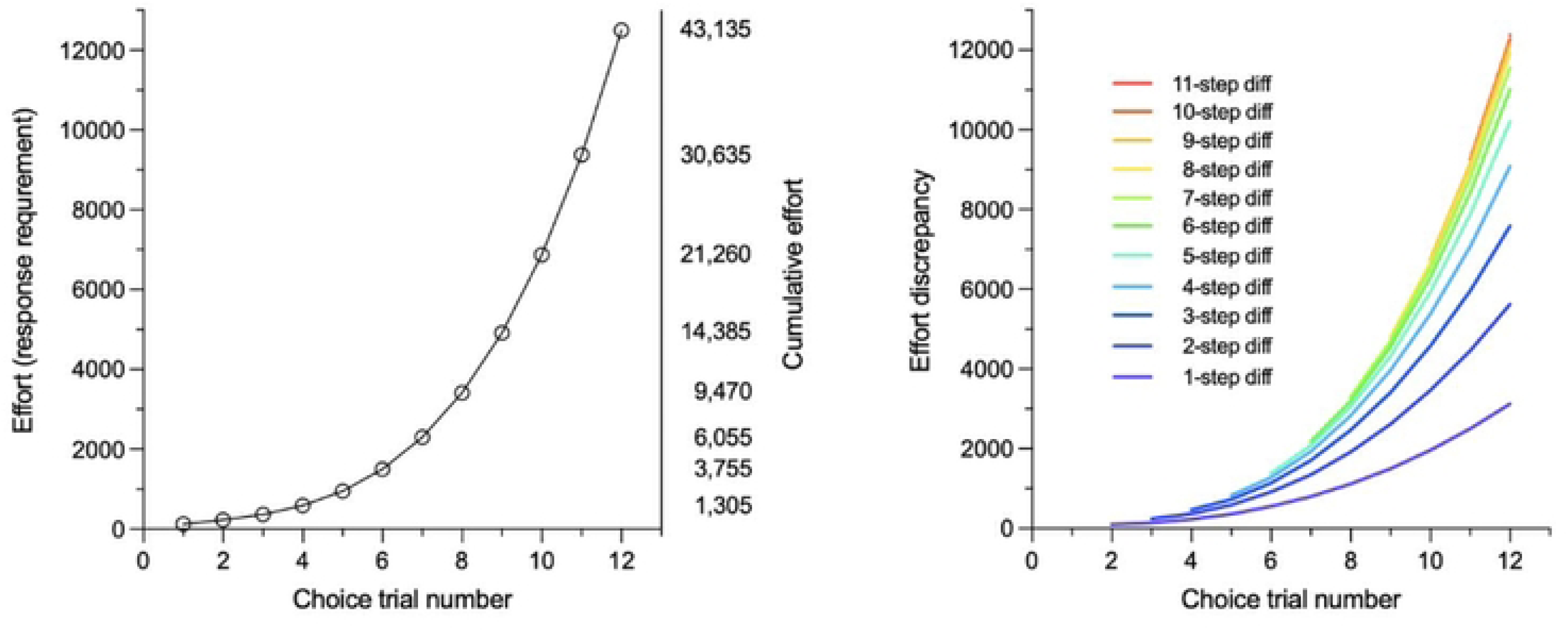
Drug/money choice progressive ratio behavioral task. *<u>Left panel</u>*: Behavioral effort (number of computer mouse clicks) required on each choice option (drug or money) of the 12-trial, exponentially-increasing progressive ratio schedule. The left Y-axis shows trial-specific effort requirements, and the right Y-axis shows the cumulative effort on a single choice option (i.e. breakpoint). *<u>Right panel</u>*: Effort discrepancy (number of mouse clicks) across trials as a function of step-size difference (current choice vs. previous choice) on the progressive ratio schedule.

### Outcome measures

Decisional speed (msec, from appearance of the two choice options to the first mouse click) was recorded for each choice a participant made. We computed within-subject median decisional speed for each response option (drug, money) and relative decisional speed (drug-money median latency difference) across all analyzable trials within condition. For purposes of analysis, choice latencies were Winsorized at an upper bound defined as the mean plus three standard deviations of all latency values.

We analyzed breakpoints for drug and money and the breakpoint difference; the latter indicates a participant’s relative effort for drug vs. money within a choice session [60]. The breakpoint difference score is calculated as the difference (in number of computer mouse clicks) between the total amount of work completed for drug and the amount completed for money.

Larger positive values indicate a greater effort for drug than money and larger negative values indicate greater effort for money. We also note that, because maximum session length is 3 hr, participants not only allocated their effort but also *time* to the alternative choice options.

### Computational modeling to predict choice sequence

Examination of choice-by-choice trial data revealed that participants generally tended to repeat previous choices. Across all choice sessions in which a switch occurred, 80% of choices were the same as the previous one. This pattern suggested that choice behavior is heavily influenced by a participant’s prior decision. To capture this dependency, we implemented a Markov chain framework with an emphasis on factors leading an individual to switch from choosing one commodity to the other (money -> drug or drug -> money).

**Equation 1** was formulated based on theoretical considerations to predict the probability of making the same choice as the previous one (habitual behavior). We hypothesized that choice switches would be more likely when (i) the *effort discrepancy* (i.e. difference in required number of mouse clicks between the next money and drug choice) between earning the same commodity and the other was high, and (ii) when participants hesitated to make a choice (longer decisional speed).

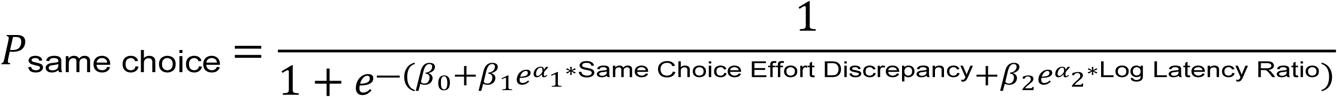

**Fig 1 (right panel)** illustrates various effort discrepancies in the context of the choice progressive ratio schedule, which follow an exponentially increasing function as more choices (steps) are repeated on the same option. As such, model inputs include the same-choice effort discrepancy (the difference in effort needed to earn the same commodity on a subsequent choice and the effort required to earn the other commodity on a subsequent choice) and the log latency ratio (the logarithm of the ratio of the decisional speed for the current choice to the previous choice). Inputs were standardized to facilitate interpretation of equation coefficients.

Output values from the probability function approaching one indicate a high likelihood of repeating the same choice and values approaching zero indicate a high likelihood of switching choices. Using this modeling approach, it was feasible only to analyze the final 10 choices made by participants during the 12-choice trial session using Equation 1, because latency information was indeterminable for the first choice made by the participant (due to uncontrolled factors at the start of each session).

The model used two free parameters, *β*_1_ and *β*_2_. These parameters were determined by fitting the probability function to each participant’s observed behavior during a choice session. Parameter fitting was accomplished using the Limited-memory Broyden–Fletcher–Goldfarb– Shanno algorithm with bound constraints [61], implemented in the scipy.optimize.minimize function in Python. Initial parameter values of *β*_1_ and *β*_2_ were selected to be zero, reflecting no contribution from the same-choice effort discrepancy faced by the participant on a choice and the observed log latency ratio, respectively; *β*_1_ and *β*_2_ were bounded between 0 and –1. More negative values indicate a greater effect of the respective inputs on switching choices.

Fixed parameters in the model included the intercept β_0_ and nonlinear scaling parameters α_1_ and α_2_. The intercept *β*_0_ captures the participant’s baseline bias toward repeating a choice. Participants repeated the majority of the choices they made. Of all analyzable choice trials, 80% were the same as the previous-trial choice. As such, *β*_0_ was bound to 4, corresponding to a high predicted baseline repetition probability of approximately 98.2% when all other coefficients equaled 0. The scaling parameters α₁ and α₂ were set to 2. Based on iterative testing, allowing *β*₀, α₁, or α₂ to vary freely did not meaningfully improve prediction accuracy, so these parameters were fixed to reduce model complexity and minimize risk of overfitting.

## Results

### Choice latency differences by participant characteristics

Relative decisional speed to choose drug vs. money (across trials within session) varied by participants’ pre-experimental cocaine use (**Fig 2, upper panels**). In Condition 2, recent cocaine users were slower to choose money than HYD (i.e. more negative median drug-money choice latencies), compared to cocaine non-users. This association was not observed in Conditions 1 or 3. In Condition 1, comparison of relative drug and money decisional latency by cocaine use was limited by small subgroups (only three cocaine non-users made both drug and money choices). In Condition 3, the $4 money alternative (higher than in the other conditions) likely increased valuation of the money reward, increasing decisional speed for money.

**Fig 2.**
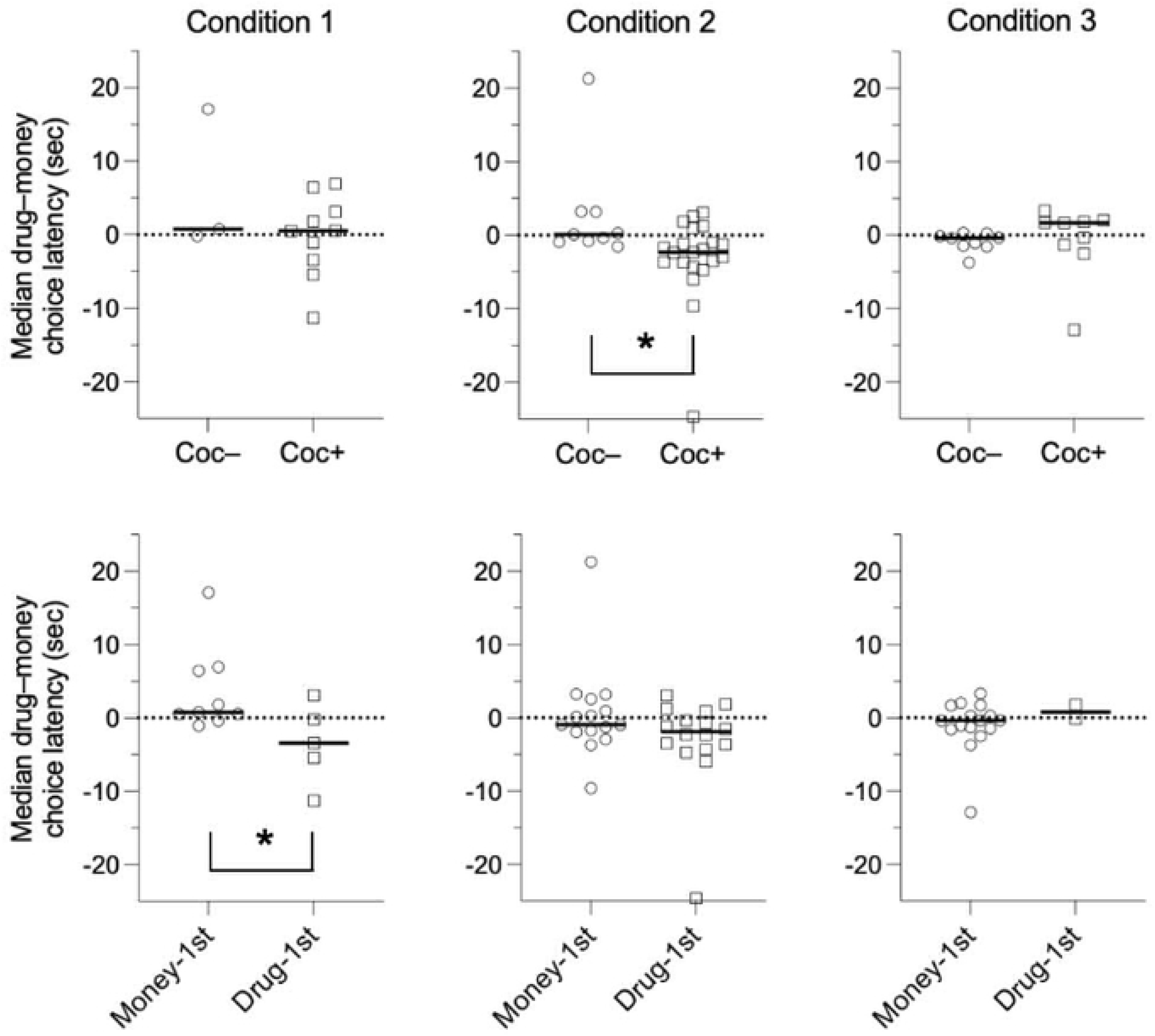
Median drug–money choice latencies, by condition and group. *<u>Upper panels</u>*: Recent cocaine users (Coc+) in Condition 2 ($2 vs. 2-mg hydromorphone, *p*=.006) took significantly longer to make choices for money than drug, compared to cocaine non-users (Coc–), whereas the groups did not differ in Condition 1 ($1 vs. 2-mg hydromorphone; *p*=.66) or Condition 3 ($4 vs. 2-mg hydromorphone, *p*=.25). An outlier at 80 sec in Condition 1 was excluded from the plot for clearer visualization. *<u>Lower panels</u>*: Drug-first responders in Condition 1 took significantly longer to make choices for money relative to drug, compared to money-first responders (*p*=.040), whereas the groups did not differ in Condition 2 (*p*=.20) or Condition 3 (*p*=.33). In each panel, the dashed line reflects the median of scattered points. *P*-values were obtained with Mann-Whitney statistical tests.

Relative decisional speed also varied between participants who made different first-trial choices for drug or money (**Fig 2, lower panels**). In Condition 1, drug-first responders were slower to choose money than HYD (i.e. more negative median drug-money choice latencies), compared to money-first responders. In Condition 2 (when the HYD dose was higher than in Condition 1 [2-mg vs. 1-mg, respectively), a drug-first vs. money-first group difference in relative decisional speed was not observed. In Condition 3, only two participants were drug-first responders, attributable to the efficacy of the larger magnitude money reinforcer in decreasing drug choices. This small subgroup size of drug-first responders in Condition 3 limited comparison of choice latencies with money-first responders.

More negative median drug-money choice latencies by recent cocaine users and drug-first responders appear to be driven by prolonged decisional latencies to money choices, rather than expedited decision-making for drug. Specifically, both recent cocaine users compared to non-users (**Fig 3, upper panels**) and drug-first compared to money-first responders (**Fig 3, lower panels**) in Conditions 1 and 2 demonstrated significantly longer median money choice latencies. (Recent cocaine use and drug-first responding status were statistically independent, χ^2^=1.50, *p*=0.22). The larger-magnitude money alternative of $4 in Condition 3 may have offset this effect (compared to $2 in Conditions 1 and 2). No discrepancies in median latencies to drug choices between drug- and money-first responders and those with and without prior cocaine use were observed in any conditions.

**Fig 3.**
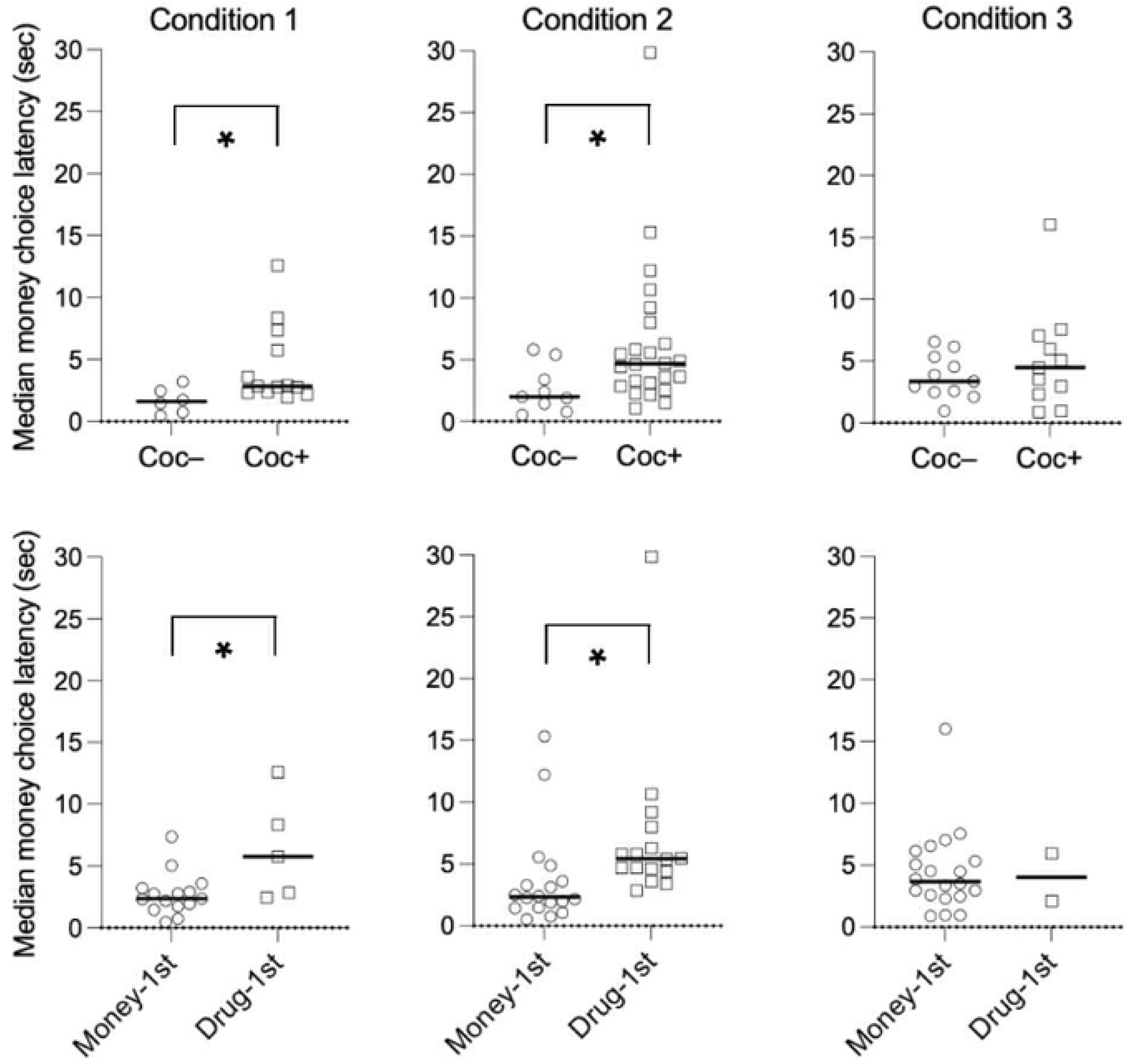
Median money choice latencies, by condition and group. *<u>Upper panels</u>*: Recent cocaine users in Condition 1 ($2 vs. 1-mg hydromorphone, *p*=.017) and Condition 2 ($2 vs. 2-mg hydromorphone, *p*=.016) took significantly longer to make choices for money, compared to cocaine non-users, whereas the groups did not differ in Condition 3 ($4 vs. 2-mg hydromorphone, *p*=.60). *<u>Lower panels</u>*: Drug-first responders in Condition 1 (*p*=.033) and Condition 2 (*p*=.001) took significantly longer to make choices for money, compared to money-first responders, whereas the groups did not differ in Condition 3 (*p*=.87). In each panel, the dashed line reflects the median of scattered points. *P*-values were obtained with Mann-Whitney statistical tests.

We observed no differences between males and females in relative decisional speeds to drug and money choices in Condition 2 (*n*s=23 vs. 8). Sex comparisons were limited by sample size in Conditions 1 and 3 as few female participants made choices for both drug and money (2 and 3, respectively).

### Faster relative drug vs. money choice latency correlates with more effortful drug seeking

Across all three experimental conditions, median drug–money choice latency difference scores inversely correlated with drug–money breakpoint difference scores (**Fig 4**). Overall variance accounted for (*r*^2^) of linear regressions between drug–money latency difference and breakpoint difference for Conditions 1, 2, and 3 were 0.41, 0.42, and 0.35, respectively. These findings suggest that, independent of experimental condition, individuals who more rapidly chose HYD relative to money expended greater effort to earn drug during choice sessions. In Conditions 1 and 2, drug-first responders’ curves were shifted upward (greater relative drug effort) relative to money-first responders.

**Fig 4.**
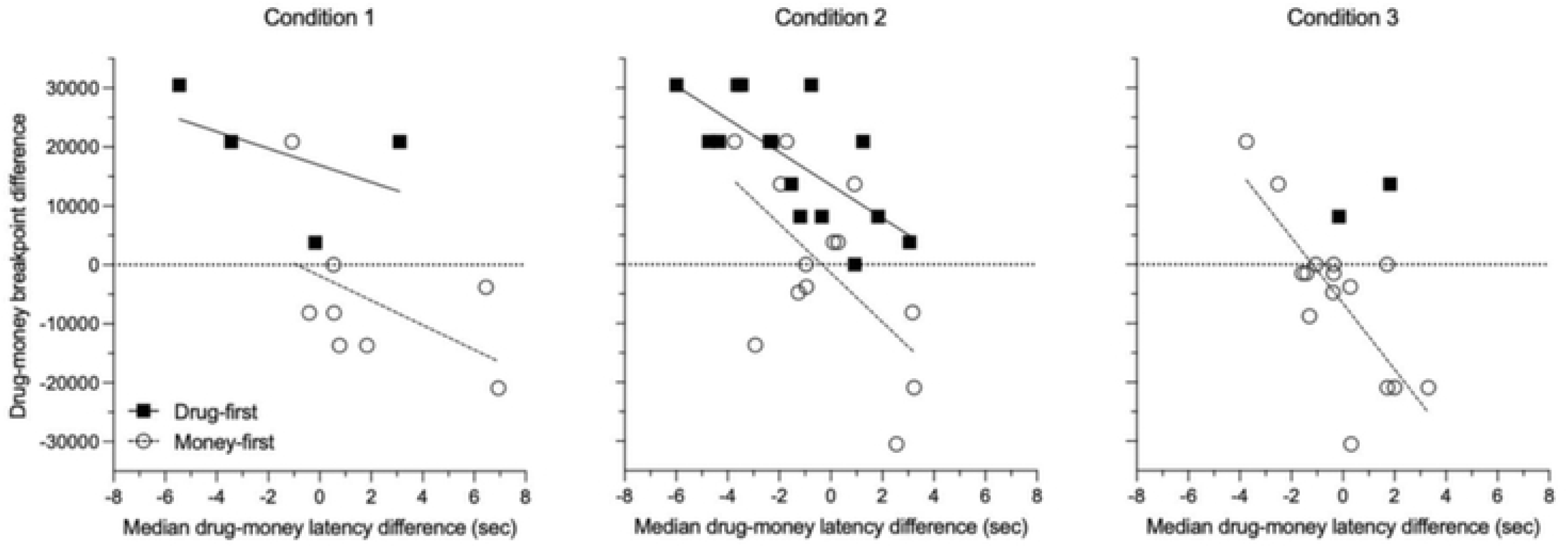
Relationship between median drug–money latency difference and drug–money breakpoint difference, by condition and group. The drug–money choice latency difference (X-axis) inversely correlated with the drug– money breakpoint difference (Y-axis) in each condition. Differences in drug and money latencies that fell outside 1.5*IQR were excluded from regression analysis. Drug-first responders (black squares) worked more for drug than money-first responders (open circles) in all groups (*p*<.001 in Conditions 1 and 2; *p*=0.025 in Condition 3 with Mann-Whitney tests). The association between median choice latency difference and breakpoint difference values was statistically significant for both drug-first and money-first subgroups in Condition 2 (*r*^2^=0.51 and 0.35), and for the money-first subgroup in Condition 3 (*r*^2^=0.59). Due to only two drug-first subjects in Condition 3, no regression line is drawn for that subgroup.

### Drug-first responders work harder to earn drug

**Fig 5** illustrates that drug-first responders made significantly more consecutive drug choices than money-first responders (Mann-Whitney *p*<0.001 for Conditions 1 and 2, and *p*=0.025 for Condition 3). The association between median drug–money latency difference and drug–money breakpoint difference values was statistically significant for both drug-first and money-first responders in Condition 2 (*r*^2^=0.51 and 0.35), and for the money-first responders in Condition 3 (*r*^2^=0.59) (**Fig 4**). Significant group differences in breakpoint difference scores and consecutive drug choices were not observed between participants who were positive vs. negative for recent cocaine use.

**Fig 5.**
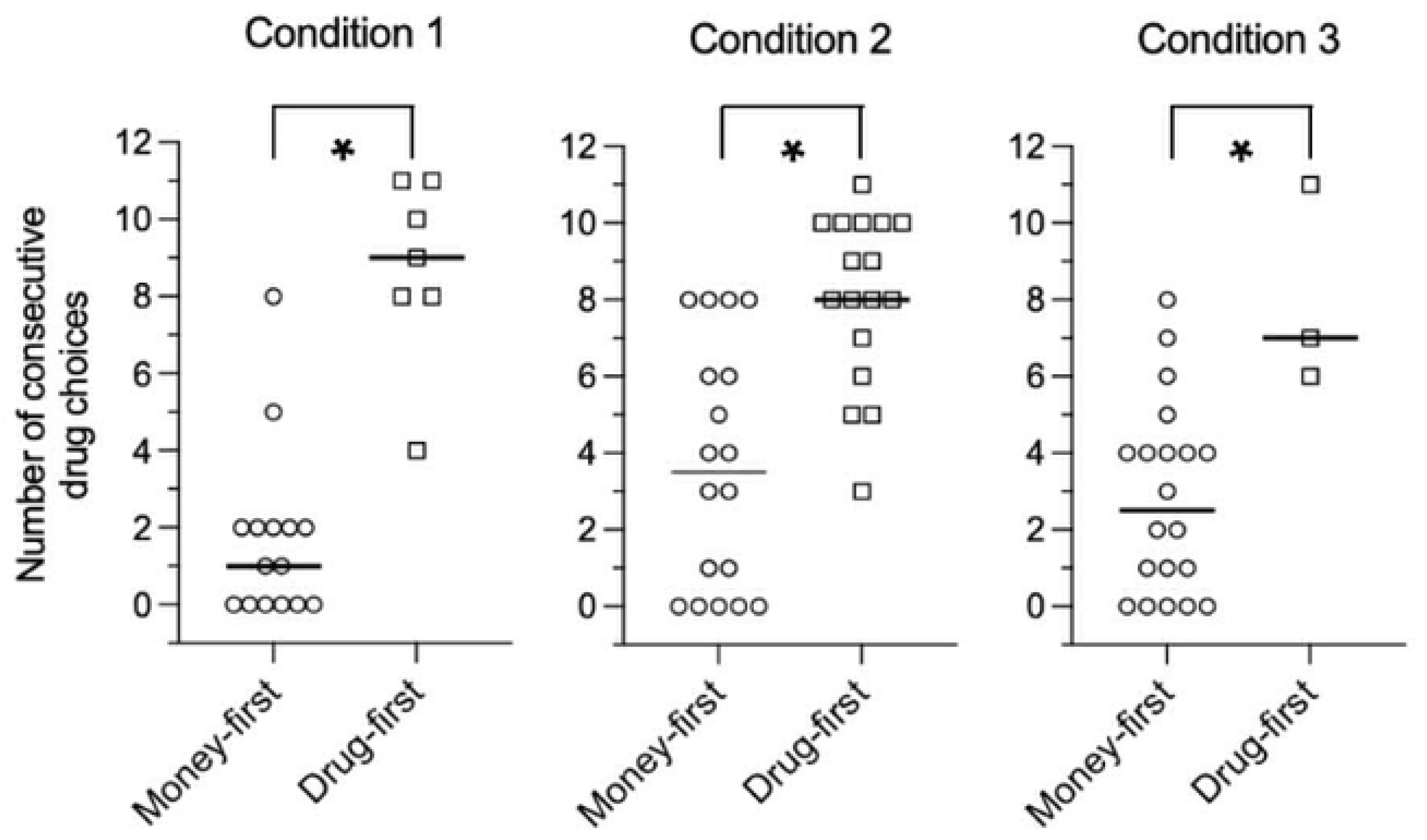
Number of consecutive drug choices in each experimental condition, stratified by money-first and drug-first subgroups. In each condition, drug-first responders made significantly more consecutive drug choices than money-first responders; Mann-Whitney *p*<0.001 for Condition 1 ($1 vs. 2-mg hydromorphone) and Condition 2 ($2 vs. 2-mg hydromorphone), and *p*=0.025 for Condition 3 ($4 vs. 2-mg hydromorphone).

**Fig 6** integrates the analyses above, using a 3D plot that depicts associations between drug–money choice latency difference, number of consecutive drug choices, and drug–money breakpoint difference, separately coded by experimental condition. Across conditions, there were associations between more consecutive drug choices, shorter latencies to choose drug than money, and a larger drug–money breakpoint difference (effort discrepancy).

**Fig 6.**
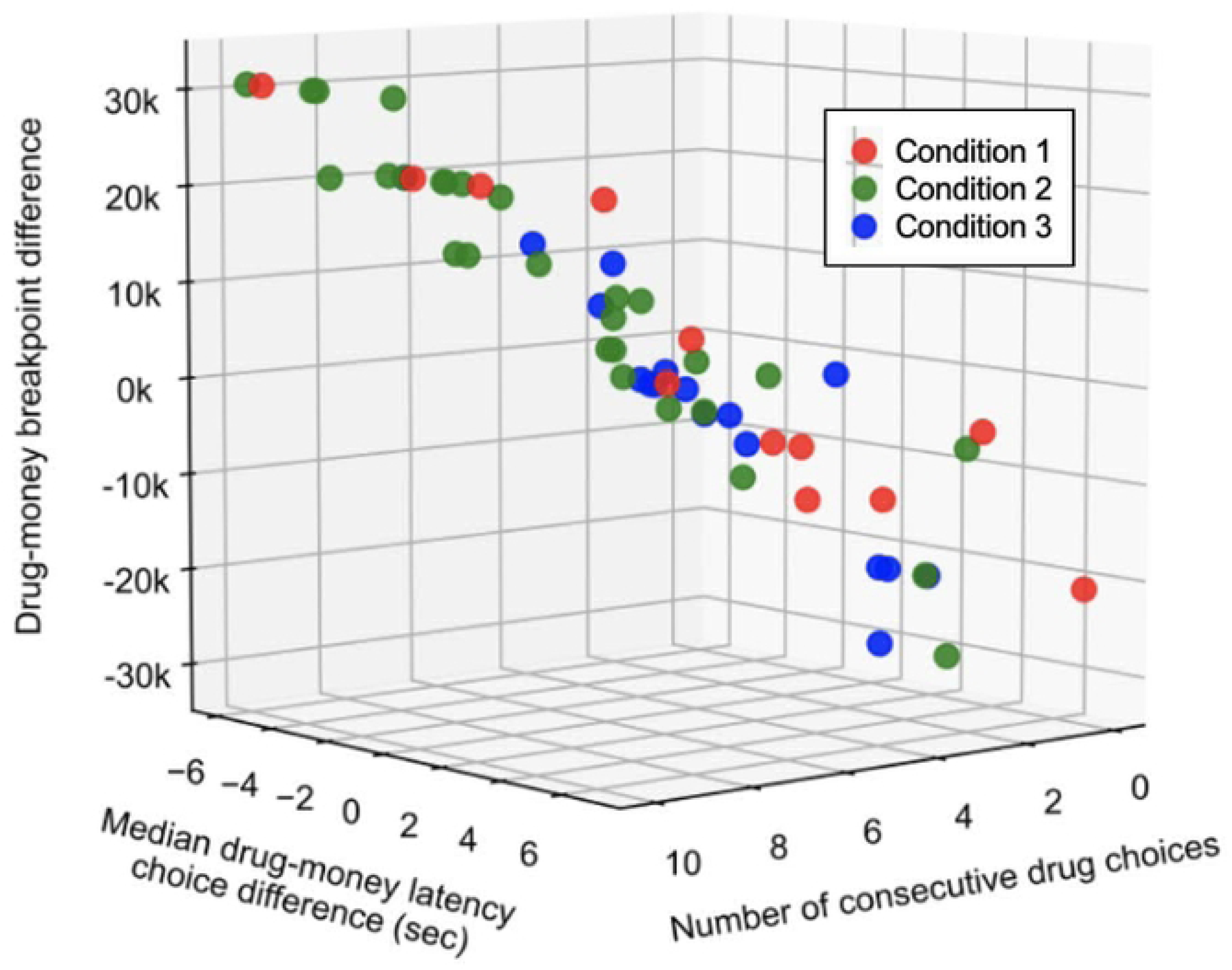
Associations between drug–money choice latency differences, consecutive drug choices, and drug–money breakpoint differences. Data are coded by Condition 1 ($1 vs. 2-mg hydromorphone; red), Condition 2 ($2 vs. 2-mg hydromorphone; green), and Condition 3 ($4 vs. 2-mg hydromorphone; blue). Across conditions, there were strong positive associations between shorter latencies to choose drug vs. money, more consecutive drug choices, and larger drug–money breakpoint difference scores.

### Model prediction of choice sequences

Across conditions, there were 64 sessions in which a participant made a choice switch for which models could be generated by fitting Equation 1. Seven of these 64 models (11%) never predicted a choice switch (i.e. model output value was never <0.5); these models were excluded from analysis as they corresponded with near-zero *β*_1_ and *β*_2_ values and lacked predictive utility. Accordingly, 57 conditions/models were analyzed across Condition 1 (*n*=15), Condition 2 (*n*=27), and Condition 3 (*n*=15).

To assess for multicollinearity between inputs, a Pearson correlation was calculated between the standardized model inputs (same-choice effort discrepancy and log latency ratio) across all data for models included in analysis. Although statistically significant, the correlation was small (*r*=0.159), suggesting negligible shared variance between predictors and minimal concern for collinearity.

**Fig 7** presents model performance for individual participants in each condition. In all but one case, models either outperformed or were comparable to a baseline persistence heuristic, in which participants were assumed to simply repeat their previous choice. Across all analyzable choice trials, participants repeated their previous choice on 80% of occasions. Fitting behavior to Eq.1 resulted in 93% prediction accuracy overall, indicating that same-choice effort and latency discrepancies explained a meaningful proportion of variance in choices.

**Fig 7.**
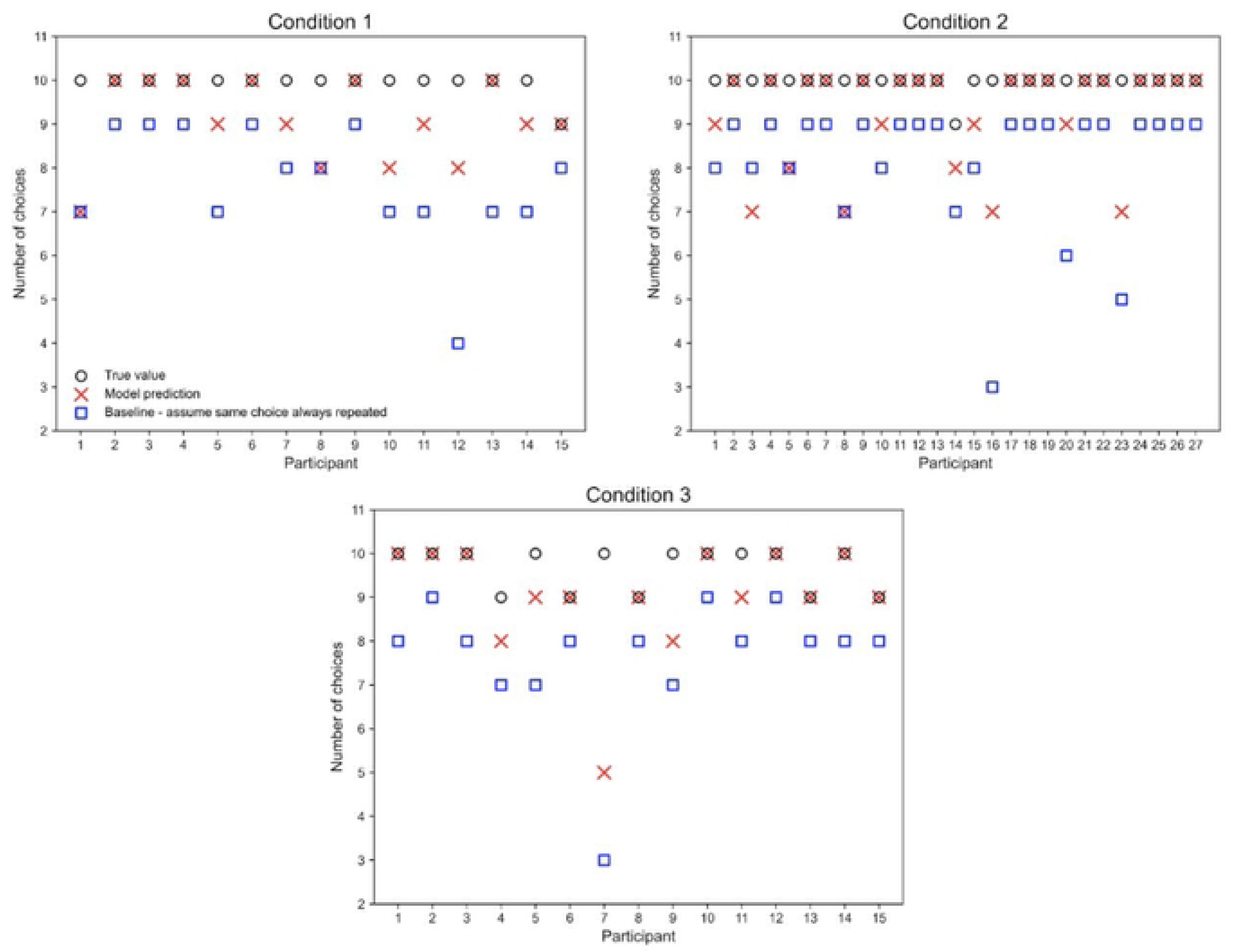
Observed choices and modeled predictions, by condition. For each participant and condition, the number of choices (regardless of drug vs. money) correctly predicted by the model (red x) and by a persistence baseline (blue squares), in which choices are assumed to repeat the previous selection, are shown relative to the total number of predictable choices (open circles). Fitted models generally outperformed or matched the baseline. Equation 1 fit behavior with 93% overall accuracy.

### Between-subject differences in model coefficients

Within each experimental condition, we compared mean ± SEM model coefficients for money-first vs. drug-first responders, adjusting for baseline characteristics: sex (male vs. female) and recent cocaine use (positive or negative). **Fig 8 (left panel)** shows that, in Condition 2, money-first responders (*n*=13) displayed a significantly more negative *β*_2_ value than money-first responders (*n*=14), −.260 ± .095 vs. −.096 ± .071, Mann-Whitney *p*=0.022. This indicates that a longer discrepancy in latencies between subsequent choices was more predictive of a choice switch in money-first than drug-first responders. Conversely, when drug-first responders hesitated during choice, they tended to make the same choice more often than money-first responders. In Condition 2, there was no money-first vs. drug-first group difference in the *β*_1_ coefficient (−.152 ± .042 vs. −.187 ± .032, respectively), suggesting no group difference to perceived effort discrepancy.

**Fig 8.**
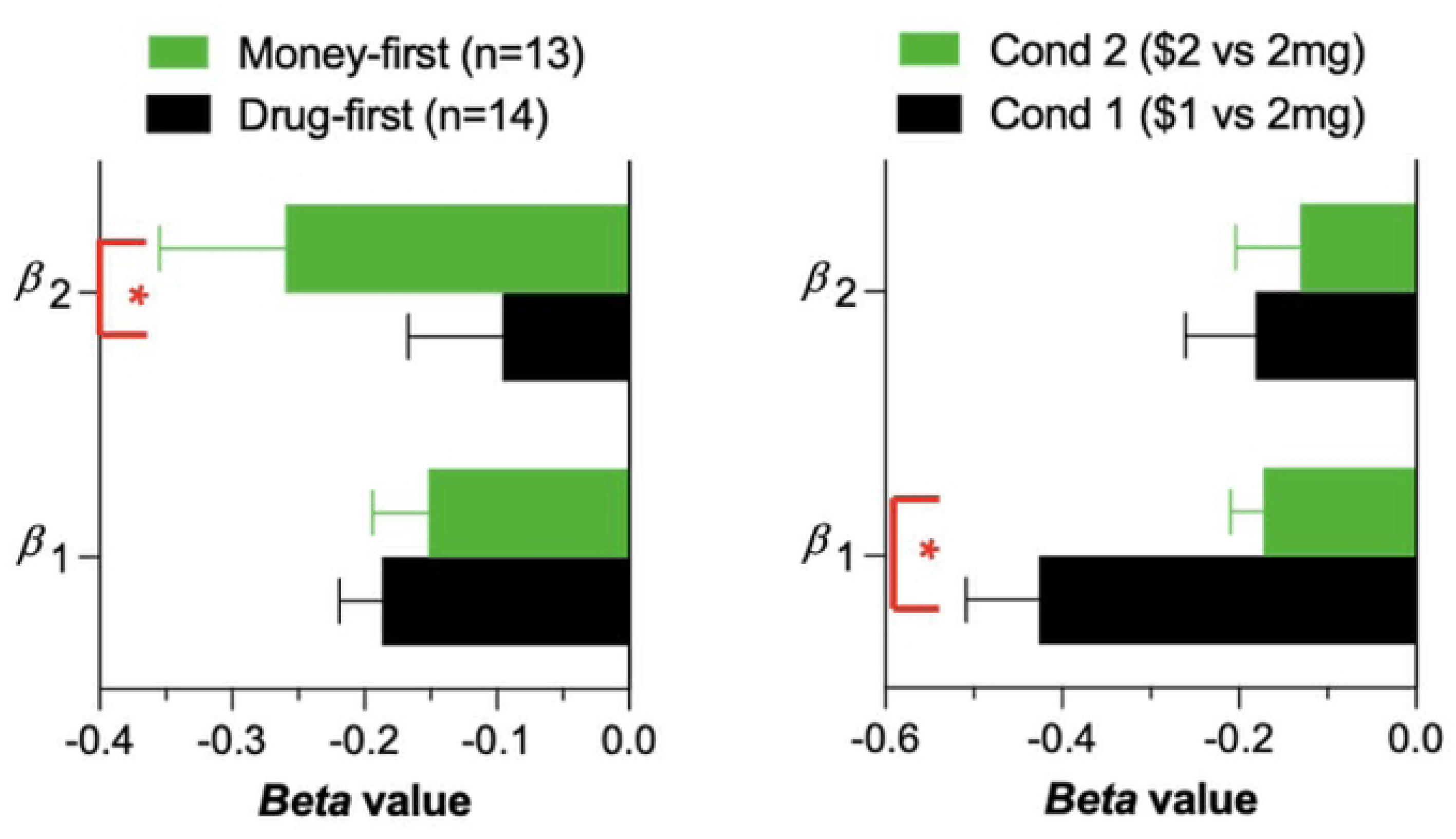
Model *β*_1_ and *β*_2_ weights, stratified by group. *<u>Left panel</u>*: Between-subject difference in model coefficients for Condition 2 ($2 vs. 2-mg hydromorphone) for money-first (*n*=13) and drug-first (*n*=14) subgroups. The *β*_2_ coefficient was significantly more negative for money-first than drug-first responders (−0.260 vs. −0.096, respectively), indicating that a larger discrepancy in successive-trial choice latencies predicted greater switching in money-first responders. There was no group difference in the *β*_1_ coefficient. *<u>Right panel</u>*: Within-subject difference in model coefficients for participants who completed Conditions 1 and 2 (*n*=14). The *β*_1_ coefficient significantly increased (became less negative) from Condition 1 to Condition 2 (−0.427 to −0.173); this indicates that as the money amount increased (at a constant hydromorphone dose), participants were less likely to switch choices when faced with greater effort discrepancies. There was no group difference in the *β*_2_ coefficient.

In Conditions 1 and 3, comparisons of coefficients between drug- and money-first responders were limited by smaller sample sizes. Only 10 and 5 models for money- and drug-first responders in Condition 1 were available for comparison, and only a single model for a drug-first responder was generated in Condition 3. Significant differences in fitted coefficients were not observed by sex or cocaine use in any experimental condition. Comparisons by sex were limited by few female participants in Conditions 1 and 3. In Condition 1, only 3 non-cocaine users were present, also limiting comparisons.

### Within-subject differences in model coefficients

Wilcoxon signed-rank tests were conducted to examine how different HYD and money reinforcer amounts affected within-subject choice behavior. **Fig 8 (right panel)** shows that *β*_1_ increased from −.427 ± .082 in Condition 1 ($2 vs. 1 mg HYD) to −.173 ± .037 in Condition 2 ($2 vs. 2 mg HYD) (*p*=0.020, *n*=14), indicating that participants became less likely to switch choices when faced with greater effort discrepancies as the HYD dose alternative increased from 1 to 2 mg (controlling for money amount). *β*_2_ did not vary between conditions (−.182 ± .079 vs. −.131 ± .073 in Conditions 1 and 2 respectively), suggesting the role of latency discrepancies on choice switching was consistent for a participant across conditions. No significant differences in any of the fitted coefficients was observed between Conditions 2 and 3; however, analysis was limited by small sample size (*n*=6).

**Figs 9** and **10** depict choice probability distributions for the only two participants who completed all three experimental conditions. Within each participant, model fits varied across conditions, illustrating context-dependent shifts in decision dynamics; however, the pattern of these shifts differed between individuals.

**Fig 9.**
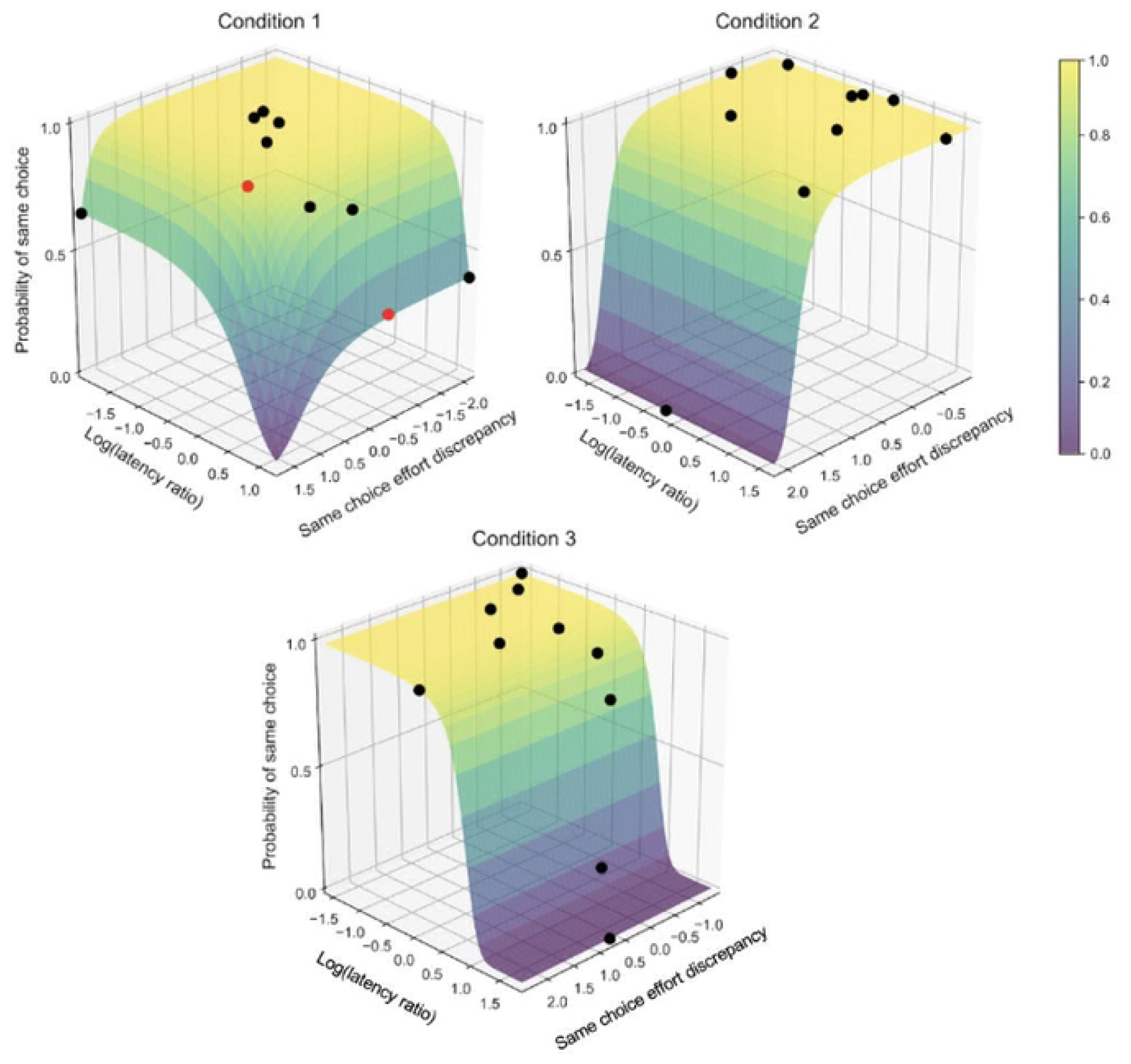
Sample probability distributions predicting the likelihood of repeating a choice for one participant who completed all three conditions. Red dots indicate the choice was incorrectly predicted; black dots indicate correctly predicted choice. In Condition 1 ($2 vs. 1-mg hydromorphone), choice switching was driven by same-choice effort and successive-choice latency discrepancies: *β*_0_ = 4.0, *β*_1_ = −0.1, *β*_2_ = −0.4. In Condition 2 ($2 vs. 2-mg hydromorphone), choice switching was driven by same-choice effort discrepancy: *β*_0_ = 4.0, *β*_1_ = −0.12, *β*_2_ = 0. In Condition 3 ($4 vs. 2-mg hydromorphone), choice switching was driven by successive-choice latency discrepancies: *β*_0_ = 4.0, *β*_1_ = 0, *β*_2_ = −0.8.

**Fig 10.**
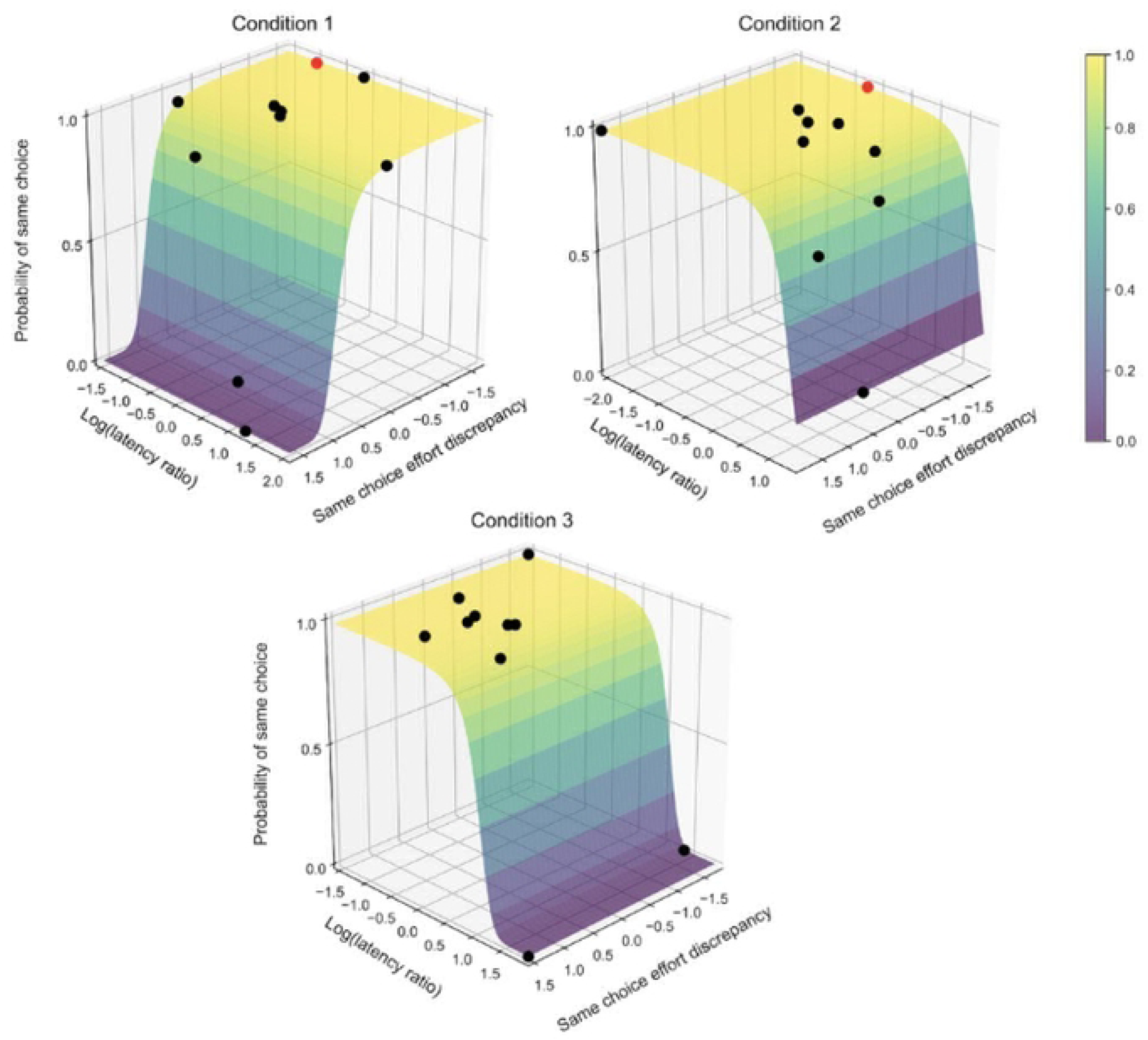
Sample probability distributions predicting the likelihood of repeating a choice for a second participant who completed all three conditions. Red dots indicate that the choice was incorrectly predicted; black dots indicate correctly predicted choices. In Condition 1 ($2 vs. 1-mg hydromorphone), choice switching was driven by same-choice effort discrepancy: *β*_0_ = 4.0, *β*_1_ = −0.71, *β*_2_ = 0. In Condition 2 ($2 vs. 2-mg hydromorphone), choice switching was driven by successive-choice latency discrepancies: *β*_0_ = 4.0, *β*_1_ = 0, *β*_2_ = −0.34. In Condition 3 ($4 vs. 2-mg hydromorphone), choice switching was driven by successive-choice latency discrepancies: *β*_0_ = 4.0, *β*_1_ = 0, *β*_2_ = −0.49.

For the first participant (**Figure 9**), choice switching in Condition 1 was influenced by both effort and latency discrepancies. In Condition 2, when HYD dose increased from 1-mg to 2-mg, switching was primarily driven by effort discrepancy. In Condition 3, when the monetary reward increased from $2 to $4, latency discrepancy emerged as the dominant predictor of switching behavior. The second participant (**Fig 10**) demonstrated a different pattern. In Condition 1, switching was primarily driven by effort discrepancy, whereas in Conditions 2 and 3, latency discrepancy was the primary predictor of switching.

Notably, both participants were male and recent cocaine users; however, the second participant made money-first choices across all conditions, whereas the first participant made a money-first choice in Condition 1 and drug-first choices in Conditions 2 and 3, suggesting that initial choice behavior may contribute to individual differences in model dynamics.

## Discussion

When a person chooses to pursue and take psychoactive drugs such as opioids, they temporarily reduce or forego consumption of other reinforcers; repetitive drug-seeking prolongs this opportunity cost for natural reinforcers and is the hallmark of addiction. The present analysis uses computational methods to characterize and predict the behavior of chronic drug users who, under experimentally varying conditions, were given the opportunity to choose between and, through effortful seeking, earn alternative rewards of different magnitudes. Non-treatment seeking, regular heroin users stabilized on buprenorphine were able to work for hydromorphone and/or money amounts during a choice task with exponentially-increasing effort requirements across trials to earn the same commodity. As we reported in each of the four source studies contributing data to this analysis [56–59], opioid withdrawal symptoms were suppressed to negligible levels during buprenorphine 8-mg/day stabilization and participants were selected to be free of major affective disturbance (e.g. anxiety or depressive disorders); thus, choices were presumably not motivated by negative reinforcement (which could overshadow effects of other parameters). Accordingly, the present computational model involves competing positive reinforcement between an opioid vs. nondrug choice alternative, and differential motivation where discrepancy in choice latency and effortful behavior can be measured.

We found that individual differences in prior-month cocaine use, first-trial drug choices, and reinforcer magnitudes for drug and money strongly influenced relative decisional speeds for HYD. In Condition 2, recent cocaine users exhibited longer decisional speeds for money ($2) relative to HYD (2 mg), suggesting that cocaine/heroin polysubstance users devalue non-drug rewards compared to heroin-only users. This pattern was not observed in Condition 3, in which the money amount was doubled to $4, increasing its salience compared to the 2 mg HYD alternative. Across experimental conditions, longer decisional latencies for money relative to drug correlated with increased effortful drug seeking. When examined more closely, in Conditions 1 and 2, both recent cocaine users and drug-first responders demonstrated longer decisional latencies specifically when making *money* choices compared to those without recent cocaine use and money-first responders, respectively. In contrast, there were no significant group differences in decisional latencies for *drug* choices, which suggests *delayed engagement with alternative nondrug reinforcers* may influence drug seeking. These results highlight the utility of programming environmental alternatives to drug use, and against the view that addiction is immutable or compulsive.

Initial choice for HYD or money in a choice session strong predicted subsequent (i.e. later trials within-session) effortful drug seeking. Drug-first responders made more consecutive drug choices (longer response chains) and exerted more effort to obtain drug, suggesting that early decisions reflect a drug-oriented motivational state. In drug vs. nondrug choice models, the first choice should be considered salient because it presumably reflects the subject’s latent preferred strategy or goal-directedness in that specific context (25, 27, 40, 43, 50, 62). Our results suggests that first-trial choice may be a proxy for willingness to expend effortful behavior on that option. This finding is also consistent with the pico-economic phenomenon of choice ‘bundling’ or ‘pre-commitment’ (49). Notably, increasing the value of the money alternative reduced the likelihood of making a drug-first choice, which again (as with response latencies) suggests that providing attractive alternative rewards can be effective in curbing drug choice and seeking behavior. This parametric variation in monetary amount was intended as an analog of contingency management, in which nondrug incentives have generally been shown to reduce drug use (63, 64) and improve retention (65) during OUD treatment.

Using a theory-driven Markov chain modeling approach, within-subject trial-by-trial choices were predicted with high accuracy (93%) using fluctuations in effort requirements and decisional speed. Differences in decision latency between subsequent choices was less correlated with choice-switching behavior among participants who made a first choice for HYD compared to those who first chose money. This could indicate that individuals who are biased toward seeking drug, as indicated by their first choice (when all other session conditions are equal), are less prone to internal deliberation and may instead rely more on automatic/habitual information processing when making decisions.

The relative reinforcing value of two concurrent alternatives (e.g. drug vs. nondrug), can dynamically shift within a session depending on response contingencies (e.g. differential effort required to earn one vs. the other reinforcer, leading to devaluation/discounting of the ‘more expensive’ option). Notably, we found that participants who completed both Conditions 1 and 2 demonstrated significantly less sensitivity to effort discrepancy in Condition 2, which offered a larger HYD dose alternative of 2-mg relative to 1-mg in Condition 1. This indicates a greater willingness to repeat choices, even when doing so required more work when the drug alternative is more salient.

Two participants completed all three experimental conditions. Interestingly, for both, their choice behavior was not universally driven by same-choice effort discrepancy (*β*_1_) or successive-choice latency discrepancies (*β*_2_) but, instead, differed across conditions. While this observation is obviously limited to these two participants, and must be cautiously interpreted, this pattern is consistent with the idea that environmental contingencies can shape choices differently across individuals. The Markov computational approach used here allowed for person-specific model fitting; future studies should consider individual difference factors might modify computational outcomes.

The present study has several limitations. First, although statistical power was enhanced by pooling participants across studies that shared identical experimental conditions, some subgroup comparisons were constrained by small sample sizes. For instance, the number of female participants was low in each condition, limiting our ability to assess sex differences in decision-making behavior; this is an important topic for future research. Second, while we varied HYD dose and money amounts across studies, the range of values was not large, which may limit generalizability. Third, our modeling approach was restricted to dynamic parameters, due to the limited number of analyzable trials per session and to reduce the risk of overfitting. While this yielded accurate predictions of trial-by-trial choices, the model likely does not fully capture the complexity of the decision-making process. Additionally, changes in choice latency between subsequent choices may not only reflect internal deliberative processes. Although latency is often used as a proxy for decisional difficulty, it may be confounded by external factors in our experimental paradigm as participants had unlimited time to choose on each trial. Consequently, extended latencies between choices could also reflect factors such as fatigue, disengagement, or distraction, rather than hesitation or decisional conflict. Fourth, although Markov modeling was an effective approach for the present data, other computational modeling methods could be explored. For instance, diffusion modeling is another computational approach that can be used to predict binary outcomes from latency to choice, among other inputs. This type of modeling typically applies to paradigms in which subjects make a large number of choices and typically involve forced, rapid decisions to best isolate underlying cognitive dynamics (Voss et al., 2013). Fifth, we did not measure the potential association between session-baseline opioid craving and drug choice/seeking; it is possible that higher craving could alter one or more model parameters such as β_0_ (i.e. baseline bias toward repeating choices) or β_1_ (same-choice effort discrepancy).

Sixth, we examined drug choice/seeking in the absence of opioid withdrawal, which has a strong motivational impact (which we elected to control for here). Different modeling results would be expected in the absence of buprenorphine maintenance, so this remains an area for future study. Seventh, these studies were conducted prior to the current wave of high-potency synthetic opioid use. It is therefore unclear whether the current modeling results might generalize to users of fentanyl or other similar opioids, which would also be important to investigate. Finally, while prior-month cocaine use influenced our findings, other non-opioid substance use or psychiatric comorbidities (which were minimized in these studies through exclusion criteria) are clinically relevant and might also affect our understanding of context-dependent choices. Despite these limitations, this innovative modeling approach accounted for a significant proportion of choice behavior and could be a useful tool for identifying underlying processes in decision-making.

## Conclusions

To our knowledge, this is the first study to provide a computational model of dynamic trial-by-trial drug-seeking (and reinforced) behavior under experimentally controlled conditions with individuals who are chronic regular (i.e. habitual) drug users. Our findings suggest that drug seeking is influenced by initial choices, decisional speed, and alternative reward magnitudes.

Furthermore, this work demonstrates the feasibility of using theory-driven computational modeling to investigate the underlying psychological processes driving drug-seeking behavior.

## Data Availability

De-identified data for this secondary analysis are publicly available at: https://github.com/Mark-Greenwald-WSU/Computational-modeling-of-opioid-seeking

## Acknowledgments

The authors thank all participants; Dr. Leslie Lundahl for overseeing psychiatric screening of participants; Dr. Caren Steinmiller for research coordination; Ken Bates for recruitment; and Lark Cederlind, Debra Kish, Joi Moore, Lisa Sulkowski, and Melissa Williams for assisting with data collection and management.

## Author Contributions (CRediT)

**Amolak Jhand**: Conceptualization, Formal analysis, Software, Visualization, Writing – original draft and editing. **Mark Greenwald**: Funding acquisition, Project administration, Conceptualization, Methodology, Supervision, Data curation, Validation, Visualization, Formal analysis, Writing – original draft & editing.

## Ethics Statement

All source studies with human participants were reviewed and approved by the local IRB. All participants provided written informed consent to participate in the study. All methods were performed in accordance with relevant guidelines and regulations.

